# Sexual violence, PrEP discussion, and PrEP use among HIV-negative men who have sex with men in 23 U.S. urban areas – National HIV Behavioral Surveillance, 2017

**DOI:** 10.1101/2023.10.04.23296565

**Authors:** Jincong Q. Freeman, Johanna Chapin-Bardales, Susan Cha, Cyprian Wejnert, Amy R. Baugher, the NHBS Study Group

**Affiliations:** Oak Ridge Institute for Science and Education, Oak Ridge, TN, USA; Division of HIV Prevention, National Center for HIV, Viral Hepatitis, STD, and TB Prevention, Centers for Disease Control and Prevention, Atlanta, GA, USA

**Keywords:** sexual violence, PrEP discussion, PrEP use, men who have sex with men, National HIV Behavioral Surveillance

## Abstract

**Background:** Men who have sex with men (MSM) who experience sexual violence are at increased risk for HIV. Pre-exposure prophylaxis (PrEP) is effective in preventing HIV infection. Associations between sexual violence and PrEP discussion or PrEP use among MSM are not well-understood.

**Methods:** National HIV Behavioral Surveillance used venue-based sampling methods to recruit and interview MSM in 23 U.S. urban areas in 2017. We estimated the prevalence of sexual violence and examined associations between sexual violence and PrEP discussion with a health care provider (HCP) or PrEP use among HIV-negative MSM in the past 12 months. We reported weighted percentages and 95% confidence intervals (CI). Adjusted prevalence ratios (aPR) with 95% CIs were calculated using logistic regression with predicted margins to compare groups.

**Results:** Among 7,121 HIV-negative MSM, 4.2% (95% CI: 3.6%-4.8%) experienced sexual violence in the past 12 months. Sexual violence was not independently associated with PrEP discussion with HCP (47.6% vs. 40.0%; aPR = 1.16, 95% CI: 0.98-1.37). MSM who experienced sexual violence were more likely to use PrEP than those who did not experience sexual violence, even after adjusting for demographic differences (34.9% vs. 25.7%; aPR = 1.34, 95% CI: 1.07-1.67).

**Conclusions:** Overall PrEP discussion and PrEP use were low among HIV-negative MSM. PrEP use was higher among MSM who experienced sexual violence. Supportive patient-provider relationships that foster PrEP discussion and sexual violence screening in healthcare settings may be important to identifying HIV risk and PrEP needs while assessing MSM’s safety.

## Introduction

Sexual violence is a serious public health issue among gay, bisexual, and other men who have sex with men (MSM) in the United States. The National Intimate Partner and Sexual Violence Survey reported that 40% of gay men and 47% of bisexual men had experienced sexual violence (i.e., being made to penetrate someone else, sexual coercion, unwanted sexual contact, and non-contact unwanted sexual experiences) in their lifetime (Walters et al., 2013). Further, experience of sexual violence has been associated with adverse health outcomes such as depression and HIV infection (Buller et al., 2014; Houston & McKirnan, 2007)(Braitstein et al., 2006). MSM in the U.S. are disproportionately affected by HIV, accounting for 65% of new HIV diagnoses in 2019 (*Centers for Disease Control and Prevention. HIV Surveillance Report, 2019; vol.32*, May 2021). A meta-analysis has demonstrated that sexual violence perpetrated by an intimate partner is associated with greater odds of having HIV among MSM (Buller et al., 2014).

Oral pre-exposure prophylaxis (PrEP) is a highly effective antiretroviral regimen against HIV infection when taken as prescribed (Chou et al., 2019; Grant et al., 2010; Spinner et al., 2016). With increasing PrEP awareness and acceptability among MSM (Holloway et al., 2020; Sullivan et al., 2020), routine PrEP discussions with health care providers are essential. Without such discussions, providers may miss opportunities to screen for PrEP or offer PrEP to their patients, including MSM who experience sexual violence. PrEP may be particularly useful to MSM who experience sexual violence, as an HIV prevention tool that does not rely on sex partners’ condom use and can be controlled by the individual. PrEP use has increased dramatically among MSM since first approved by the U.S. Food and Drug Administration in 2012 (*U.S. Food and Drug Administration. Truvada for PrEP Fact Sheet: Ensuring Safe and Proper Use*, 2012); however, overall PrEP uptake remains suboptimal among MSM (Finlayson et al., 2019; Sullivan et al., 2020).

Prevalence of PrEP discussion and PrEP use among MSM who experience sexual violence, and associations between sexual violence experience and PrEP discussion or PrEP use among MSM are not well-understood. A recent study has indicated that MSM who experience forced sex perpetrated by an intimate partner are less likely to use PrEP (Braksmajer et al., 2020). Understanding the association between sexual violence experience and PrEP discussion or PrEP use among MSM may help inform violence and HIV prevention and PrEP interventions for MSM and training for PrEP providers. In this analysis, we investigated the prevalence of sexual violence in the past 12 months and examined associations between recent sexual violence experience and PrEP discussion with a health care provider or PrEP use among a multisite sample of HIV-negative MSM.

## Methods

### National HIV Behavioral Surveillance

The Centers for Disease Control and Prevention’s (CDC) National HIV Behavioral Surveillance (NHBS) conducts rotating, annual, bio-behavioral surveillance among three key populations: MSM, persons who inject drugs, and heterosexually active persons at increased risk for HIV (Gallagher et al., 2007). Data for this analysis were collected in 2017 using venue-based, time-space sampling (VBS) methods to recruit and interview MSM in 23 U.S. metropolitan statistical areas (MSAs) with high HIV burden: Atlanta, Georgia; Baltimore, Maryland; Boston, Massachusetts; Chicago, Illinois; Dallas, Texas; Denver, Colorado; Detroit, Michigan; Houston, Texas; Los Angeles, California; Memphis, Tennessee; Miami, Florida; Nassau-Suffolk Counties, New York; New Orleans, Louisiana; New York City, New York; Newark, New Jersey; Philadelphia, Pennsylvania; Portland, Oregon; San Diego, California; San Francisco, California; San Juan, Puerto Rico; Seattle, Washington; Virginia Beach, Virginia; and Washington, DC (*Centers for Disease Control and Prevention. HIV Infection Risk, Prevention, and Testing Behaviors Among Men Who Have Sex With Men-National HIV Behavioral Surveillance, 23 U.S. Cities, 2017*, 2019; MacKellar et al., 2007). Eligible participants were offered a standardized questionnaire and HIV testing (MacKellar et al., 2007). Eligibility criteria for NHBS participation included being assigned as male sex at birth and self-identifying as a man, being at least 18 years old, living in a participating MSA, ever having had sex with another man, not having previously participated in the 2017 NHBS, being able to provide informed consent, and being able to complete the questionnaire in English or Spanish. This analysis is further limited to MSM who were sexually active (defined as having had sex with at least one male partner in the past 12 months) and who received a negative NHBS HIV test result. NHBS 2017 data were weighted to account for unequal selection probabilities, multiplicity, and nonresponse, allowing us to produce representative estimates for venue-attending MSM in the 23 MSAs. NHBS sampling methods and procedures have been previously published (*Centers for Disease Control and Prevention. HIV Infection Risk, Prevention, and Testing Behaviors Among Men Who Have Sex With Men-National HIV Behavioral Surveillance, 23 U.S. Cities, 2017*, 2019). NHBS was determined by the CDC to be public health surveillance. Local institutional review boards in each of the 23 participating MSAs approved NHBS activities (*Centers for Disease Control and Prevention. Guidelines for Defining Public Health Research and Public Health Non-Research Revised October 4, 1999*, 1999; *United States Department of Health and Human Services. Protection of Human Subjects. CFR 45, Part 46. Revised January 2009*, 2009). All NHBS participants provided informed consent.

### Measures

Recent sexual violence experience, the main independent variable of interest, was measured by asking participants if anybody forced or pressured them to have oral or anal sex when they did not want to in the past 12 months (yes/no). The outcomes of interest were PrEP discussion with a health care provider and PrEP use in the past 12 months. To assess PrEP discussion, we asked participants, “In the past 12 months, have you had a discussion with a health care provider about taking PrEP?” If participants were aware of PrEP, had visited a provider in the past 12 months, and discussed PrEP with a provider in the past 12 months, they were categorized as having discussed PrEP with a health care provider. To assess PrEP use, we asked participants, “In the past 12 months, have you taken PrEP to reduce the risk of getting HIV?” If participants were aware of PrEP and reported having taken PrEP in the past 12 months, they were categorized as having used PrEP.

Demographic and behavioral characteristics included age, race/ethnicity, educational attainment, health insurance status, employment status, census region, condomless anal sex, number of male sex partners, exchange sex, last male sex partner’s HIV status, HIV testing offered by a health care provider, likely having an indication for PrEP, and PrEP awareness. Hispanic or Latino MSM were of any race. Condomless anal sex was defined as having had condomless anal sex with a male partner in the past 12 months. Exchange sex was defined as having either received or given money or drugs in exchange for sex with a male partner in the past 12 months. Last sex partner’s HIV status was assessed by asking participants if they had known their last male sex partner’s HIV status and if so, what their last sex partner’s HIV status was the last time they had sex. Participants aware of PrEP reported having ever heard of PrEP. Participants were considered to have been offered HIV testing by a health care provider if they had visited a provider and a provider offered an HIV test in the past 12 months. Likely having an indication for PrEP was defined as having had either a male HIV-positive partner or ≥2 male sex partners in the past 12 months; and having had either a self-reported sexually transmitted infection or condomless anal sex with a male partner in the past 12 months (*Preexposure Prophylaxis for the Prevention of HIV Infection in the United States—2017 Update: A Clinical Practice Guideline.*, 2017).

### Statistical Analysis

We estimated the prevalence of sexual violence in the past 12 months among HIV-negative MSM and described their demographic and behavioral characteristics overall and by recent sexual violence experience. Weighted percentages or median with 95% confidence intervals (CI) were reported; *p*-values were calculated using Rao-Scott chi-square tests for categorical data and Wilcoxon rank-sum tests for discrete data. We examined associations between recent sexual violence experience and PrEP discussion with a health care provider or PrEP use in the past 12 months among HIV-negative MSM. Unadjusted prevalence ratios (PR) and adjusted prevalence ratios (aPR) with 95% CIs were calculated using logistic regression with predicted margins (Bieler et al., 2010). Adjusted models were controlled for age, race/ethnicity, educational attainment, health insurance status, employment status, and census region. We did not report categories with an unstable coefficient of variation (CV≥0.3) and excluded them from logistic regression models due to sparse data. *P*-values of <0.05 were considered statistically significant. We performed all analyses and accounted for complex survey design and weights using PROC SURVEYMEANS and PROC SURVEYFREQ in SAS 9.4 (SAS Institute Inc., Cary, NC) and PROC RLOGIST in SAS-callable SUDAAN 11.0 (RTI International Research, Research Triangle Park, NC).

## Results

In total, 7,121 sexually active HIV-negative MSM were included in this analysis. Of these, 4.2% (95% CI: 3.6%-4.8%) experienced sexual violence in the past 12 months. Demographic and behavioral characteristics are presented in Table 1. Overall, the largest percentages of MSM were 30 years of age or older (39.8%, 95% CI: 38.1%-41.6%) and were White (27.3%, 95% CI: 25.7%-28.9%). Most MSM had education beyond high school (59.4%, 95% CI: 57.6%-61.2%), were insured (60.2%, 95% CI: 58.4%-62.0%), or were employed either full-time or part-time (60.7%, 95% CI: 58.9%-62.4%). More than half of MSM (53.5%, 95% CI: 51.8%-55.3%) had condomless anal sex with a male partner in the past 12 months (Table 1).

**Table 1.** Characteristics of HIV-negative MSM by recent sexual violence experience in 23 U.S. urban areas – NHBS, 2017.

|  | Total (N=7,121) |  | Experienced Sexual Violence in the Past 12 Months (n=460) |  | Did Not Experience Sexual Violence in the Past 12 Months (n=6,661) |  | *p*-value<sup>†</sup> |
| --- | --- | --- | --- | --- | --- | --- | --- |
|  | N* | %* (95% CI) | n* | %* (95% CI) | n* | %* (95% CI) |  |
| **Age group (years)** |  |  |  |  |  |  |  |
| 18-29 | 3,063 | 33.6 (31.7-35.5) | 269 | 59.3 (52.6-65.9) | 2,794 | 44.9 (42.6-47.2) | <0.0001 |
| ≥30 | 4,058 | 39.8 (38.1-41.6) | 191 | 40.7 (34.1-47.4) | 3,867 | 55.1 (52.8-57.4) |  |
| **Race/Ethnicity\*** |  |  |  |  |  |  |  |
| Hispanic or Latino | 2,001 | 25.1 (23.5-26.7) | 103 | 26.4 (20.1-32.8) | 1,898 | 34.7 (32.7-36.7) | <0.0001 |
| American Indian or Alaska Native | 49 | 0.7 (0.4-1.0) | —§ | —§ | 41 | 0.8 (0.5-1.1) |  |
| Asian | 192 | 2.2 (1.8-2.6) | —§ | —§ | 182 | 3.1 (2.5-3.7) |  |
| Black | 1,670 | 14.6 (13.2-16.0) | 105 | 17.0 (11.8-22.3) | 1,565 | 20.1 (18.1-22.0) |  |
| Native Hawaiian or Other Pacific Islander | 32 | 0.3 (0.1-0.4) | —§ | —§ | —§ | —§ |  |
| White | 2,773 | 27.3 (25.7-28.9) | 192 | 43.2 (36.2-50.3) | 2,581 | 36.8 (34.8-38.9) |  |
| Multiple races | 365 | 3.2 (2.7-3.7) | 37 | 7.5 (4.0-11.0) | 328 | 4.2 (3.5-4.9) |  |
| **Highest level of education** |  |  |  |  |  |  |  |
| High school/GED or less | 1,479 | 14.0 (12.8-15.2) | 100 | 18.3 (13.0-23.6) | 1,379 | 19.1 (17.5-20.7) | 0.7709 |
| Beyond high school | 5,640 | 59.4 (57.6-61.2) | 360 | 81.7 (76.4-87.0) | 5,280 | 80.9 (79.3-82.5) |  |
| **Health insurance status** |  |  |  |  |  |  |  |
| Yes | 5,819 | 60.2 (58.4-62.0) | 374 | 84.3 (79.2-89.5) | 5,445 | 81.9 (80.4-83.5) | 0.4024 |
| No | 1,291 | 13.1 (12.1-14.2) | 85 | 15.7 (10.5-20.8) | 1,206 | 18.1 (16.5-19.6) |  |
| **Employment status** |  |  |  |  |  |  |  |
| Unemployed | 1,420 | 12.7 (11.7-13.8) | 129 | 29.0 (22.9-35.1) | 1,291 | 16.6 (15.2-18.0) | <0.0001 |
| Full- or part-time employed | 5,700 | 60.7 (58.9-62.4) | 331 | 71.0 (64.9-77.1) | 5,369 | 83.4 (82.0-84.8) |  |
| **Census region** |  |  |  |  |  |  |  |
| Midwest | 606 | 3.4 (2.7-4.2) | 48 | 4.1 (1.9-6.4) | 558 | 4.7 (3.6-5.8) | 0.0117 |
| Northeast | 1,287 | 14.0 (12.4-15.6) | 81 | 17.0 (11.6-22.5) | 1,206 | 19.2 (17.1-21.4) |  |
| South | 2,655 | 27.3 (25.2-29.5) | 137 | 32.1 (25.2-39.0) | 2,518 | 37.6 (34.6-40.5) |  |
| West | 2,326 | 27.1 (24.8-29.5) | 188 | 46.4 (39.1-53.6) | 2,138 | 36.4 (33.3-39.4) |  |
| Puerto Rico | 247 | 1.5 (1.0-1.9) | —§ | —§ | 241 | 2.1 (1.5-2.8) |  |
| **Condomless anal sex with a male partner in the past 12 months** |  |  |  |  |  |  |  |
| Yes | 5,111 | 53.5 (51.8-55.3) | 367 | 77.4 (71.4-83.4) | 4,744 | 72.4 (70.7-74.1) | 0.1309 |
| No | 2,001 | 20.2 (18.9-21.4) | 92 | 22.6 (16.6-28.6) | 1,909 | 27.6 (25.9-29.3) |  |
| **Number of male sex partners in the past 12 months, median (95% CI)** | 3.9 (3.6-4.1) |  | 8.8 (7.1-10.5) |  | 3.7 (3.5-3.9) |  | <0.0001 |
| **Exchange sex with a male partner in the past 12 months<sup>¶</sup>** |  |  |  |  |  |  |  |
| Yes | 566 | 5.1 (4.4-5.8) | 100 | 20.6 (15.1-26.0) | 466 | 6.1 (5.2-7.0) | <0.0001 |
| No | 6,553 | 68.6 (66.9-70.4) | 359 | 79.4 (74.0-84.9) | 6,194 | 93.9 (93.0-94.8) |  |
| **Last male sex partner's HIV status** |  |  |  |  |  |  |  |
| HIV-negative | 4,323 | 48.3 (46.5-50.1) | 251 | 57.6 (51.0-64.2) | 4,072 | 63.1 (61.3-64.9) | 0.1255 |
| HIV-positive | 407 | 4.2 (3.6-4.8) | 27 | 4.5 (1.8-7.1) | 380 | 5.6 (4.7-6.4) |  |
| Don't know | 2,346 | 24.4 (23.1-25.7) | 177 | 37.9 (31.4-44.5) | 2,169 | 31.3 (29.6-33.0) |  |

Abbreviations: MSM, men who have sex with men; NHBS, National HIV Behavioral Surveillance; CI, confidence interval; GED, general educational development.
\*Frequencies are unweighted, and percentages are weighted.
<sup>†</sup>P-values were calculated using Rao-Scott chi-square tests for categorical data and Wilcoxon rank-sum tests for discrete data.
<sup>‡</sup>Hispanic or Latinos men can be of any race.
<sup>§</sup>Categories with an unstable coefficient of variation ( $CV \geq 0.3$ ) were not reported.
<sup>¶</sup>Exchange sex with a male partner is defined as having received or given money or drugs in exchange for having sex with a male partner in the past 12 months.

By recent sexual violence experience (Table 1), MSM who experienced sexual violence reported higher percentages of being aged 18-29 years (59.3%, 95% CI: 52.6%-65.9%), being unemployed (29.0%, 95% CI: 22.9%-35.1%), and having had exchange sex with a male partner in the past 12 months (20.6%, 95% CI: 15.1%-26.0%) than those who did not experience sexual violence. Compared to MSM who did not experience sexual violence (median = 3.7, 95% CI: 3.5-3.9), those who experienced sexual violence reported having had a greater number of male sex partners (median = 8.8, 95% CI: 7.1-10.5) in the past 12 months.

In Figure 1, the majority of MSM who experienced sexual violence were aware of PrEP (87.9%, 95% CI: 83.4%-92.5%), likely had an indication for PrEP (81.6%, 95% CI: 75.6%-87.5%), and were offered HIV testing by a health care provider in the past 12 months (56.0%, 95% CI: 48.7%-63.3%), and results were not significantly different from those who did not experience sexual violence.

**Figure 1.**
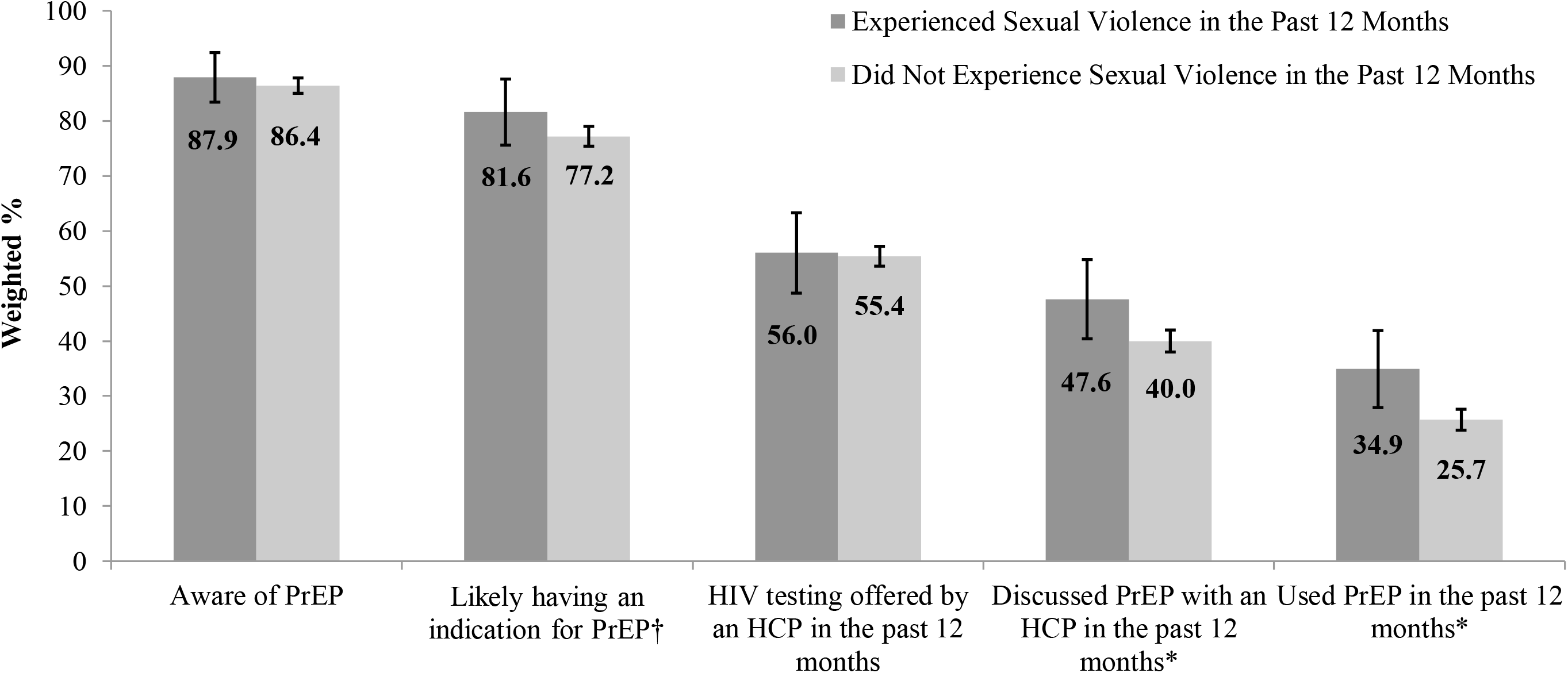
PrEP-related characteristics of HIV-negative MSM by recent sexual violence experience in 23 U.S. urban areas – NHBS, 2017

### PrEP discussion

Among MSM who experienced sexual violence, 47.6% (95% CI: 40.4%-54.7%) had discussed PrEP with a health care provider in the past 12 months, compared to 40.0% (95% CI: 38.0%-42.1%) among those who did not experience sexual violence (Figure 1). After adjusting for demographic differences, the association between recent sexual violence experience and PrEP discussion was not significant (aPR = 1.16, 95% CI: 0.98-1.37) (Table 2).

**Table 2.** Associations between recent sexual violence experience and PrEP discussion or PrEP use among HIV-negative MSM in 23 U.S. urban areas – NHBS, 2017.

|  | **Discussed PrEP with a Health Care Provider in the Past 12 Months** |  | **Did Not Discuss PrEP with a Health Care Provider in the Past 12 Months** |  |  |  |  |  |
| --- | --- | --- | --- | --- | --- | --- | --- | --- |
|  | n* | Row %* (95% CI) | n* | Row %* (95% CI) | PR (95% CI) | p-value | aPR<sup>†</sup> (95% CI) | p-value |
| **Sexual Violence in the Past 12 Months** |  |  |  |  |  |  |  |  |
| Total | 2,793 | 38.0 (36.1-39.8) | 4,323 | 55.9 (54.0-57.7) |  |  |  |  |
| Experienced | 215 | 47.6 (40.4-54.7) | 244 | 52.4 (45.3-59.6) | 1.19 (1.01-1.40) | 0.0477 | 1.16 (0.98-1.37) | 0.0968 |
| Did not experience | 2,578 | 40.0 (38.0-42.1) | 4,079 | 60.0 (57.9-62.0) | 1.0 (Referent) |  | 1.0 (Referent) |  |

|  | **Used PrEP in the Past 12 Months** |  | **Did Not Use PrEP in the Past 12 Months** |  |  |  |  |  |
| --- | --- | --- | --- | --- | --- | --- | --- | --- |
|  | n* | Row %* (95% CI) | n* | Row %* (95% CI) | PR (95% CI) | p-value | aPR<sup>†</sup> (95% CI) | p-value |
| **Sexual Violence in the Past 12 Months** |  |  |  |  |  |  |  |  |
| Total | 1,782 | 24.6 (22.9-26.3) | 5,335 | 69.2 (67.4-71.1) |  |  |  |  |
| Experienced | 147 | 34.9 (27.9-41.9) | 312 | 65.1 (58.1-72.1) | 1.36 (1.10-1.68) | 0.0079 | 1.34 (1.07-1.67) | 0.0150 |
| Did not experience | 1,635 | 25.7 (23.8-27.6) | 5,023 | 74.3 (72.4-76.2) | 1.0 (Referent) |  | 1.0 (Referent) |  |

Abbreviations: PrEP, pre-exposure prophylaxis; MSM, men who have sex with men; NHBS, National HIV Behavioral Surveillance; CI, confidence interval; PR, unadjusted prevalence ratio; aPR, adjusted prevalence ratio.
\*Frequencies are unweighted, and percentages are weighted.
<sup>†</sup>Adjusted for age, race/ethnicity, highest level of education, health insurance status, employment status, and census region.

### PrEP use

Among MSM who experienced sexual violence, 34.9% (95% CI: 27.9%-41.9%) had used PrEP in the past 12 months, while 25.7% (95% CI: 23.8%-27.6%) of those who did not experience sexual violence used PrEP in the past 12 months (Figure 1). MSM who experienced sexual violence were more likely than those who did not experience sexual violence to have used PrEP, even after adjusting for demographic differences (aPR = 1.34, 95% CI: 1.07-1.67).

### Discussion

Overall, a weighted prevalence of 4.2% of HIV-negative MSM experienced sexual violence in the past 12 months; most of these men were between the ages of 18-29 years. One in 5 MSM who experienced sexual violence had exchange sex with a male partner in the past 12 months. Moreover, MSM who experienced sexual violence reported a median of 9 male sex partners in the past 12 months. These results align with literature on sexual violence among MSM, which indicates that MSM who experience sexual violence are more likely than those who do not experience sexual violence to be younger, engage in exchange sex, and have multiple male sex partners (Duncan et al., 2018; Nasrullah et al., 2015). These findings are concerning because MSM are disproportionately affected by HIV (*Centers for Disease Control and Prevention. HIV Surveillance Report, 2019; vol.32*, May 2021), and those who experience sexual violence are at increased risk for HIV (Buller et al., 2014). Providers should take sexual history and behaviors into consideration when assessing the safety and HIV risk of MSM who experience sexual violence, which can help facilitate discussions of PrEP and increase PrEP uptake.

Health care providers play an important role in assessing HIV risk and facilitating access to HIV prevention tools such as PrEP. PrEP may be a particularly critical HIV prevention option for MSM who experience sexual violence given uptake and use can be directed by the individual unlike other strategies that may require negotiation with a male sex partner (e.g., condom use, positioning). We found that MSM reported high PrEP awareness regardless of recent sexual violence experiences, though a small proportion (12%-14%) reported being unaware. There was no difference in likely having an indication for PrEP between MSM who experienced sexual violence and those who did not experience sexual violence. This finding may be explained by high condomless anal sex regardless of recent sexual violence experiences given condomless anal sex was a component of our PrEP indication variable. Only half of sexually active MSM were offered HIV testing by a provider in the past 12 months, among both those who did or did not recently experience sexual violence. While many MSM receive HIV testing in non-clinical settings, this is still suboptimal given recommendations of annual HIV testing for sexually active MSM(Duncan et al., 2018; Nasrullah et al., 2015) and because offering HIV testing is a key first step for providers to engage in conversations about HIV risk and PrEP indications with their male patients, which may lead to better understanding of sexual violence experiences.

Less than half of MSM had discussed PrEP with a health care provider in the past 12 months. Although the percentage of PrEP discussion was higher among MSM who experienced sexual violence, it was not significantly different from those who did not experience sexual violence. One possible explanation could be that the prevalence of sexual violence was too small to detect differences. Alternatively, providers may not assess male patients’ recent experiences of sexual violence as a potential HIV risk factor that could inform PrEP discussion. Providers may consider directly screening MSM for experiences of sexual violence, both for identifying potential HIV risk, engaging in discussions about PrEP, and as an opportunity to refer these men to violence and HIV prevention services.

Gender stereotypes of victims of violence being women can be a barrier to men being assessed or linked to needed support and social services, or services not being sufficiently tailored to men, including MSM (Baral et al., 2015; Bates et al., 2019; Javanbakht et al., 2019). Furthermore, this analysis’ definition of sexual violence covers sexual violence from any perpetrator, which may be in the context of intimate partner relationships, other social or familial relationships, the criminal justice system, or other contexts; needed support and services could be different depending on the perpetrator. Acknowledging the importance of the patient-provider relationship and the clinical environments supporting violence and HIV prevention with MSM patients, clinics and providers should foster a stigma-free, discrimination-free, culturally competent environment where male patients, especially MSM, can feel comfortable disclosing their experiences of sexual violence and discussing HIV prevention options such as PrEP.

Approximately a quarter of MSM used PrEP in the past 12 months; one third of those who experienced sexual violence had used PrEP. We found higher prevalence of PrEP use among MSM who experienced sexual violence; however, this finding is inconsistent with a previous study that found forced sex perpetrated by an intimate partner to be negatively associated with PrEP use among MSM (Braksmajer et al., 2020). However, this analysis defines sexual violence from any perpetrator, which may help explain the difference. It is possible that experiences of sexual violence increased perceived risk for HIV infection, which led to higher PrEP use among MSM in our analysis. Prior studies have documented that MSM with high perceived HIV risk are more likely to be aware of and willing to use PrEP compared to those with low perceived HIV risk (Holt et al., 2012; Kesler et al., 2016; Underhill et al., 2018). In addition, MSM who experienced sexual violence in our analysis also reported higher percentages of having had exchange sex and greater number of male sex partners, which could also contribute to higher perceived HIV risk; this higher perceived HIV risk may have resulted in greater PrEP use even if likely having an indication for PrEP remained similar between those who did and those who did not recently experience sexual violence. Information regarding the influence of sexual behaviors and sexual violence experiences on perceived HIV risk and PrEP discussion or PrEP use among MSM are lacking and NHBS 2017 did not assess perceived HIV risk among MSM. Future research is needed to assess the effects of sexual violence experience and of sexual violence perpetration on perceived HIV risk and the role of perceived HIV risk as a barrier or a facilitator for HIV prevention such as PrEP discussion or PrEP use among MSM.

### Limitations

This analysis has several limitations. First, sexual violence was self-reported and is subject to recall bias or social desirability bias, which may have underestimated the prevalence of recent sexual violence experience. Second, temporality between experience of sexual violence and PrEP discussion or PrEP use could not be established due to the cross-sectional survey design. Third, it is possible that unmeasured factors (e.g., screening for violence, perceived HIV risk, relationship to perpetrator, PrEP acceptability) may help better explain the association between recent sexual violence experience and PrEP use among HIV-negative MSM. Lastly, the findings are only generalizable to venue-attending MSM who live in urban areas. Data were weighted to adjust for potential sampling biases; however, findings may not be generalizable to all HIV-negative MSM in the U.S.

## Conclusions

Overall PrEP discussion with a health care provider and PrEP use were low among sexually active HIV-negative MSM. The prevalence of PrEP use was higher among MSM who experienced sexual violence than those who did not experience sexual violence, though was not optimal for either group. Strategies to promote PrEP discussions between providers and their patients and to increase PrEP uptake among MSM and those who experience sexual violence are needed. Recent sexual violence experience was associated with PrEP use but was not associated with PrEP discussion among MSM. Supportive patient-provider relationships that foster discussions of PrEP and screening for sexual violence in healthcare settings may be important to identifying HIV risk and PrEP needs while assessing MSM’s safety.

## Data Availability

All data analyzed are not publicly available.

## Acknowledgements

We thank the NHBS participants, participating facilities and project area staff, and members of the NHBS study group: Atlanta, GA: Pascale Wortley, Jeff Todd, David Melton; Baltimore, MD: Colin Flynn, Danielle German; Boston, MA: Monina Klevens, Rose Doherty, Conall O’Cleirigh; Chicago, IL: Stephanie Masiello Schuette, Antonio D. Jimenez; Dallas, TX: Jonathon Poe, Margaret Vaaler, Jie Deng; Denver, CO: Alia Al-Tayyib, Melanie Mattson; Detroit, MI: Vivian Griffin, Emily Higgins, Mary-Grace Brandt; Houston, TX: Salma Khuwaja, Zaida Lopez, Paige Padgett; Los Angeles, CA: Ekow Kwa Sey, Yingbo Ma; Memphis, TN: Shanell L. McGoy, Meredith Brantley, Randi Rosack; Miami, FL: Emma Spencer, Willie Nixon, David Forrest; Nassau-Suffolk Counties, NY: Bridget Anderson, Ashley Tate, Meaghan Abrego; New Orleans, LA: William T. Robinson, Narquis Barak, Jeremy M. Beckford; New York City, NY: Sarah Braunstein, Alexis Rivera, Sidney Carrillo Newark, NJ: Barbara Bolden, Afework Wogayehu, Henry Godette; Philadelphia, PA: Kathleen A. Brady, Chrysanthus Nnumolu, Jennifer Shinefeld; Portland, OR: Sean Schafer, E. Roberto Orellana, Amisha Bhattari; San Diego, CA: Anna Flynn, Rosalinda Cano; San Francisco, CA: H. Fisher Raymond, Theresa Ick; San Juan, PR: Sandra Miranda De León, Yadira Rolón-Colón; Seattle, WA: Tom Jaenicke, Sara Glick; Virginia Beach, VA: Celestine Buyu, Toyah Reid, Karen Diepstra; Washington, DC: Jenevieve Opoku, Irene Kuo; CDC: Monica Adams, Christine Agnew Brune, Qian An, Alexandra Balaji, Dita Broz, Janet Burnett, Johanna Chapin-Bardales, Melissa Cribbin, YenTyng Chen, Paul Denning, Katherine Doyle, Teresa Finlayson, Senad Handanagic, Brooke Hoots, Wade Ivy, Kathryn Lee, Rashunda Lewis, Evelyn Olansky, Gabriela Paz-Bailey, Taylor Robbins, Catlainn Sionean, Amanda Smith, Cyprian Wejnert, Mingjing Xia.

## Notes

### Competing Interest Statement

The authors have declared no competing interest.

### Funding Statement

National HIV Behavioral Surveillance (NHBS) is funded by the Centers for Disease Control and Prevention (CDC) through a cooperative agreement with state and local health departments. This project was supported in part by
an appointment to the Research Participation Program at the CDC administered by the Oak Ridge Institute for Science and Education through an interagency agreement between the U.S. Department of Energy and the CDC. The findings and conclusions of this analysis are those of the authors and do not necessarily represent the views of the Centers for Disease Control and Prevention and the U.S. Department of Health and Human Services.

### Author Declarations

All National HIV Behavioral Surveillance (NHBS) participants provided informed consent. NHBS was determined by the Centers for Disease Control and Prevention to be a routine disease surveillance system and therefore exempted from IRB review.

## References

Baral, S. D., Friedman, M. R., Geibel, S., Rebe, K., Bozhinov, B., Diouf, D., Sabin, K., Holland, C. E., Chan, R., & Caceres, C. F. (2015, Jan 17). Male sex workers: practices, contexts, and vulnerabilities for HIV acquisition and transmission. Lancet, 385(9964), 260–273. 10.1016/S0140-6736(14)60801-1

Bates, E. A., Klement, K. R., Kaye, L. K., & Pennington, C. R. (2019). The impact of gendered stereotypes on perceptions of violence: a commentary. Sex Roles, 81, 34–43. 10.1007/s11199-019-01029-9

Bieler, G. S., Brown, G. G., Williams, R. L., & Brogan, D. J. (2010, Mar 1). Estimating model-adjusted risks, risk differences, and risk ratios from complex survey data. Am J Epidemiol, 171(5), 618–623. 10.1093/aje/kwp440

Braitstein, P., Asselin, J. J., Schilder, A., Miller, M. L., Laliberte, N., Schechter, M. T., & Hogg, R. S. (2006, Oct). Sexual violence among two populations of men at high risk of HIV infection. AIDS Care, 18(7), 681–689. 10.1080/13548500500294385

Braksmajer, A., Walters, S. M., Crean, H. F., Stephenson, R., & McMahon, J. M. (2020, Aug). Pre-exposure Prophylaxis Use Among Men Who Have Sex with Men Experiencing Partner Violence. AIDS Behav, 24(8), 2299–2306. 10.1007/s10461-020-02789-2

Buller, A. M., Devries, K. M., Howard, L. M., & Bacchus, L. J. (2014, Mar). Associations between intimate partner violence and health among men who have sex with men: a systematic review and meta-analysis. PLoS Med, 11(3), e1001609. 10.1371/journal.pmed.1001609

Centers for Disease Control and Prevention. Guidelines for Defining Public Health Research and Public Health Non-Research Revised *October 4,* 1999. (1999). Retrieved February 16 from https://www.cdc.gov/os/integrity/docs/defining-public-health-research-non-research-1999.pdf

*Centers for Disease Control and Prevention.* HIV Infection Risk, Prevention, and Testing Behaviors Among Men Who Have Sex With Men-National HIV Behavioral Surveillance, 23 U.S. Cities, 2017. (2019). Retrieved December 31 from https://www.cdc.gov/hiv/pdf/library/reports/surveillance/cdc-hiv-surveillance-special-report-number-22.pdf

Centers for Disease Control and Prevention. HIV Surveillance Report*, 2019; vol.32*. (May 2021). Retrieved June 15 from https://www.cdc.gov/hiv/library/reports/hiv-surveillance/vol-32/index.html

Chou, R., Evans, C., Hoverman, A., Sun, C., Dana, T., Bougatsos, C., Grusing, S., & Korthuis, P. T. (2019, Jun 11). Preexposure Prophylaxis for the Prevention of HIV Infection: Evidence Report and Systematic Review for the US Preventive Services Task Force. JAMA, 321(22), 2214–2230. 10.1001/jama.2019.2591

Duncan, D. T., Goedel, W. C., Stults, C. B., Brady, W. J., Brooks, F. A., Blakely, J. S., & Hagen, D. (2018, Mar). A Study of Intimate Partner Violence, Substance Abuse, and Sexual Risk Behaviors Among Gay, Bisexual, and Other Men Who Have Sex With Men in a Sample of Geosocial-Networking Smartphone Application Users. Am J Mens Health, 12(2), 292–301. 10.1177/1557988316631964

Finlayson, T., Cha, S., Xia, M., Trujillo, L., Denson, D., Prejean, J., Kanny, D., Wejnert, C., & National, H. I. V. B. S. S. G. (2019, Jul 12). Changes in HIV Preexposure Prophylaxis Awareness and Use Among Men Who Have Sex with Men - 20 Urban Areas, 2014 and 2017. MMWR Morb Mortal Wkly Rep, 68(27), 597–603. 10.15585/mmwr.mm6827a1

Gallagher, K. M., Sullivan, P. S., Lansky, A., & Onorato, I. M. (2007). Behavioral surveillance among people at risk for HIV infection in the U.S.: the National HIV Behavioral Surveillance System. Public Health Rep, 122 *Suppl 1*, 32–38. 10.1177/00333549071220S106

Grant, R. M., Lama, J. R., Anderson, P. L., McMahan, V., Liu, A. Y., Vargas, L., Goicochea, P., Casapia, M., Guanira-Carranza, J. V., Ramirez-Cardich, M. E., Montoya-Herrera, O., Fernandez, T., Veloso, V. G., Buchbinder, S. P., Chariyalertsak, S., Schechter, M., Bekker, L. G., Mayer, K. H., Kallas, E. G., Amico, K. R., Mulligan, K., Bushman, L. R., Hance, R. J., Ganoza, C., Defechereux, P., Postle, B., Wang, F., McConnell, J. J., Zheng, J. H., Lee, J., Rooney, J. F., Jaffe, H. S., Martinez, A. I., Burns, D. N., Glidden, D. V., & iPrEx Study, T. (2010, Dec 30). Preexposure chemoprophylaxis for HIV prevention in men who have sex with men. N Engl J Med, 363(27), 2587–2599. 10.1056/NEJMoa1011205

Holloway, I. W., Krueger, E. A., Meyer, I. H., Lightfoot, M., Frost, D. M., & Hammack, P. L. (2020). Longitudinal trends in PrEP familiarity, attitudes, use and discontinuation among a national probability sample of gay and bisexual men, 2016-2018. PLoS One, 15(12), e0244448. 10.1371/journal.pone.0244448

Holt, M., Murphy, D. A., Callander, D., Ellard, J., Rosengarten, M., Kippax, S. C., & de Wit, J. B. (2012, Jun). Willingness to use HIV pre-exposure prophylaxis and the likelihood of decreased condom use are both associated with unprotected anal intercourse and the perceived likelihood of becoming HIV positive among Australian gay and bisexual men. Sex Transm Infect, 88(4), 258–263. 10.1136/sextrans-2011-050312

Houston, E., & McKirnan, D. J. (2007, Sep). Intimate partner abuse among gay and bisexual men: risk correlates and health outcomes. J Urban Health, 84(5), 681–690. 10.1007/s11524-007-9188-0

Javanbakht, M., Ragsdale, A., Shoptaw, S., & Gorbach, P. M. (2019, Jun). Transactional Sex among Men Who Have Sex with Men: Differences by Substance Use and HIV Status. J Urban Health, 96(3), 429–441. 10.1007/s11524-018-0309-8

Kesler, M. A., Kaul, R., Myers, T., Liu, J., Loutfy, M., Remis, R. S., & Gesink, D. (2016, Nov). Perceived HIV risk, actual sexual HIV risk and willingness to take pre-exposure prophylaxis among men who have sex with men in Toronto, Canada. AIDS Care, 28(11), 1378–1385. 10.1080/09540121.2016.1178703

MacKellar, D. A., Gallagher, K. M., Finlayson, T., Sanchez, T., Lansky, A., & Sullivan, P. S. (2007). Surveillance of HIV risk and prevention behaviors of men who have sex with men--a national application of venue-based, time-space sampling. Public Health Rep, 122 *Suppl 1*, 39–47. 10.1177/00333549071220S107

Nasrullah, M., Oraka, E., Chavez, P. R., Valverde, E., & Dinenno, E. (2015, Aug 24). Nonvolitional sex and HIV-related sexual risk behaviours among MSM in the United States. AIDS, 29(13), 1673–1680. 10.1097/QAD.0000000000000631

Preexposure Prophylaxis for the Prevention of HIV Infection in the United States—2017 Update: A Clinical Practice Guideline. (2017). Centers for Disease Control and Prevention. Retrieved May 14 from

Spinner, C. D., Boesecke, C., Zink, A., Jessen, H., Stellbrink, H. J., Rockstroh, J. K., & Esser, S. (2016, Apr). HIV pre-exposure prophylaxis (PrEP): a review of current knowledge of oral systemic HIV PrEP in humans. Infection, 44(2), 151–158. 10.1007/s15010-015-0850-2

Sullivan, P. S., Sanchez, T. H., Zlotorzynska, M., Chandler, C. J., Sineath, R. C., Kahle, E., & Tregear, S. (2020, Mar). National trends in HIV pre-exposure prophylaxis awareness, willingness and use among United States men who have sex with men recruited online, 2013 through 2017. J Int AIDS Soc, 23(3), e25461. 10.1002/jia2.25461

U.S. Food and Drug Administration. Truvada for PrEP Fact Sheet: Ensuring Safe and Proper Use. (2012). Retrieved May 14 from https://www.fda.gov/media/83586/download

Underhill, K., Guthrie, K. M., Colleran, C., Calabrese, S. K., Operario, D., & Mayer, K. H. (2018, Oct). Temporal Fluctuations in Behavior, Perceived HIV Risk, and Willingness to Use Pre-Exposure Prophylaxis (PrEP). Arch Sex Behav, 47(7), 2109–2121. 10.1007/s10508-017-1100-8

United States Department of Health and Human Services. Protection of Human Subjects. CFR 45, Part 46*. Revised January* 2009. (2009). Retrieved February 16 from https://www.hhs.gov/ohrp/regulations-and-policy/regulations/45-cfr-46/index.html

Walters, M. L., Chen, J., & Breiding, M. J. (2013). The National Intimate Partner and Sexual Violence Survey (NISVS): 2010 Findings on Victimization by Sexual Orientation. Centers for Disease Control and Prevention. Retrieved February 24 from https://www.cdc.gov/violenceprevention/pdf/nisvs_sofindings.pdf

